# Prescription Practices of Medical Cannabinoids in Children with Cerebral Palsy - A Survey of the Swiss Cerebral Palsy Registry

**DOI:** 10.1101/2021.11.18.21266388

**Authors:** Federico Morosoli, Sandra Hunziker, Kathrin Zuercher, Anne Tscherter, Sebastian Grunt, on behalf of the Swiss Cerebral Palsy Registry Group

**Affiliations:** Division of Neuropediatrics, Development and Rehabilitation, Children’s University Hospital, Inselspital, University of Bern, Freiburgstr. 15, CH-3010Bern, Switzerland; Institute of Social and Preventive Medicine, University of Bern, Mittelstrasse 43, CH-3012 Bern, Switzerland

**Keywords:** Medical cannabis, Tetrahydrocannabinol, Cannabidiol, Cerebral palsy, Pediatrics

## Abstract

**Aim:** Medical cannabinoids are prescribed to children with cerebral palsy despite limited evidence. We aimed to assess the prescription practices of cannabinoids in children with cerebral palsy with a particular focus on indications and preparations used and how well cannabinoids are tolerated. Furthermore, we investigated how physicians acquire knowledge on cannabinoid medication.

**Methods:** We asked physicians with expertise in the care of children with CP on their prescription practices of medical cannabinoids. Data was collected through an online survey, which was distributed by email. In addition to the demographic information of participants, we also inquired about the indications for the prescription of cannabinoids, about experiences regarding efficacy and the observed side effects of the therapy.

**Results:** Seventy physicians from Europe, North America and Australia completed the survey. Forty-seven participants were experienced in the treatment of children with cerebral palsy by cannabinoids. The most common indication was epilepsy (69%), followed by spasticity (64%) and pain (63%). The prescribed preparations and doses varied considerably. Half of the participants evaluated the effect of the medical cannabinoids as moderate. Twenty-nine physicians reported side effects, most frequently in the form of drowsiness (26%), somnolence (19%), fatigue (13%), and diarrhoea (13%).

**Conclusions:** Despite the lack of evidence to date, medical cannabinoids are used to treat children with cerebral palsy in a wide variety of indications. Randomized controlled trials in this vulnerable patient group are therefore of utmost importance.

**Highlights:**

- Cannabinoids are frequently used in children with CP, despite weak evidence.
- Cannabinoids are prescribed in children with CP for different indications.
- The most common indications are epilepsy, spasticity, and pain.
- Common acute side effects are drowsiness, somnolence, fatigue, diarrhoea, and nausea.
- Long-term side effects of cannabinoids in children with CP were not observed.

## Introduction

The term cerebral palsy (CP) refers to a non-progressive disorder of posture and movement due to a non-progressive malformation or lesion in the brain [1]. A distinction is made between ataxic, dyskinetic and spastic (unilateral or bilateral) forms of CP [2]. Children with CP suffer from a variety of comorbidities in addition to motor symptoms such as epilepsy, behavioural, musculoskeletal, and nutritional issues, sleeping problems, and pain [1, 3-7]. A wide variety of therapeutic approaches and drug treatments for the individual symptom complexes of patients with CP have been described [8]. Medications mainly address the treatment of epilepsy as well as movement disorders [5, 9-11]. In addition to conventional medications, cannabinoids are increasingly used for various indications in children. Cannabis (*Cannabis sativa*) is a plant-based drug composed of over 100 phytocannabinoids. Among them, the psychoactive substance Δ9-tetrahydrocannabinol (THC) and cannabidiol (CBD) are of particular interest.

CBD has an anti-inflammatory, anti-epileptic and anxiety-alleviating effect [12] and is used in children primarily to treat epileptic seizures, anxiety disorders and psychoses [13-16]. Its use has been well studied for epilepsy. Three randomised controlled trials (RCTs) studied the effect of CBD in patients with Lennox-Gastaut syndrome and Dravet syndrome and showed a greater reduction in seizures frequency compared to placebo with good tolerance [14-17]. THC is used in children and adults for different indications [13, 18, 19]. In adults, it is mainly used for the treatment of chronic pain and spasticity in multiple sclerosis [18]. There is moderate evidence that THC reduces spasticity in adult multiple sclerosis patients [19, 20], but the evidence for the treatment of other indications in adults is weak [19]. In children, the use of THC has been described in patients with neurodegenerative diseases, traumatic brain injury, posttraumatic stress disorder and Tourette’s syndrome [13], to treat chemotherapy-induced nausea and vomiting [21] and to treat spasticity in the context of paediatric palliative care [22]. So far, there is little evidence on its effectiveness in children. In January 2019, a systematic literature review summarised the literature on the use of cannabinoids for spasticity with a special focus on children [23]. A conclusion regarding the effect of cannabinoids on childhood spasticity could not be drawn. Shortly afterwards (March 2019), an RCT on the effect of Nabiximols (Sativex®) in 72 children with spasticity (mostly with spastic CP) was published. It showed no significant improvement in spasticity 12 weeks after treatment compared to placebo [24].

Despite the limited evidence for cannabinoids in children (with exception of CBD in Lennox-Gastaut and Dravet syndrome), cannabinoids seem to be used in Switzerland and other countries for a wide range of indications in children with CP. Information on prescription practices is mostly based on anecdotal reports and case descriptions. A recent Swiss parent survey showed that THC and CBD are mainly used in children with neurological disorders [25]. It remains unclear in which context and for which indications cannabinoids are prescribed to children with CP and what experience specialists have had in treating children with CP with cannabinoids.

Therefore, we sent a questionnaire to medical experts in Europe, North America, and Australia with the aim of finding out for which indications cannabinoids were prescribed, what effects and side effects were observed, and how the prescribing physicians acquired their knowledge about treatment with cannabinoids.

## Methods

### Study design and data collection

This prospective observational study collected data from physicians who treat children (<18 years) affected by CP. A questionnaire in English was developed and reviewed by the steering board members of the Swiss Cerebral Palsy Registry. The revised questionnaire (Appendix B.1) was used for data collection through an online survey on the SurveyMonkey® platform (San Mateo, California, USA; www.surveymonkey.com). Access to the questionnaire was given by e-mail. The targeted participants were physicians in Europe, North America, and Australia with expertise in the care of children with CP. The initial address list was developed by the Steering Board of the Swiss Cerebral Palsy Registry in collaboration with the Swiss Academy of Childhood Disability and was a convenience sample of personal contacts. Addressees were invited to forward the survey to relevant colleagues. Data collection took place from October 2019 to February 2020.

The questionnaire was composed of three sections: (I) four questions to assess if the participants fulfil the inclusion criteria; (II) demographic data such as sex, age, country of residence, medical specialization, work experience, and workplace of the participants; (III) prescription practices of medical cannabinoids in children with CP, including participants years of experience on the subject, indications and formulas used, observed efficacy and short-/long-term adverse effects.

### Inclusion criteria and definition of groups

Participants fulfilled the inclusion criteria and obtained access to the complete questionnaire, if they answered positively to one or more of the questions of section (I):

- Have you already worked with patients treated with cannabinoid drugs?
- Were any of these in a paediatric age (0-18y)?
- Were any of these children/adolescents diagnosed with cerebral palsy?
- Did you ever experience a situation in which the prescription of medical cannabinoids in this type of patients was debated, but for some reason not prescribed?

Physicians who answered negatively to all four questions or who did not complete the questionnaire were excluded from the study.

Participants affirming all four questions of section (I) were defined as experienced in the treatment of children with CP using medical cannabinoids, while participants affirming one to three questions were defined as unexperienced.

### Statistical analysis

We used descriptive statistics to characterize participants by experience of medical cannabis in children with CP (yes vs. no) and by region and assessed differences between groups using chi-square, Fisher’s exact, or Wilcoxon rank-sum tests. All analyses were done in Stata (version 15.1, College Station, TX, USA).

### Ethical statement

For this study, no approval by an institutional review board was required within the context of Swiss law.

## Results

The total number of physicians responding to the survey was 96. We excluded 7 physicians for not completing the survey and 19 physicians for not meeting at least one of the four inclusion criteria. Hence, 70 participants were included in the study (Figure 1). Of these, 23 (33%) belonged to the unexperienced group and 47 (67%) were experienced in the prescription of medical cannabinoids to children with CP.

**Figure 1:**
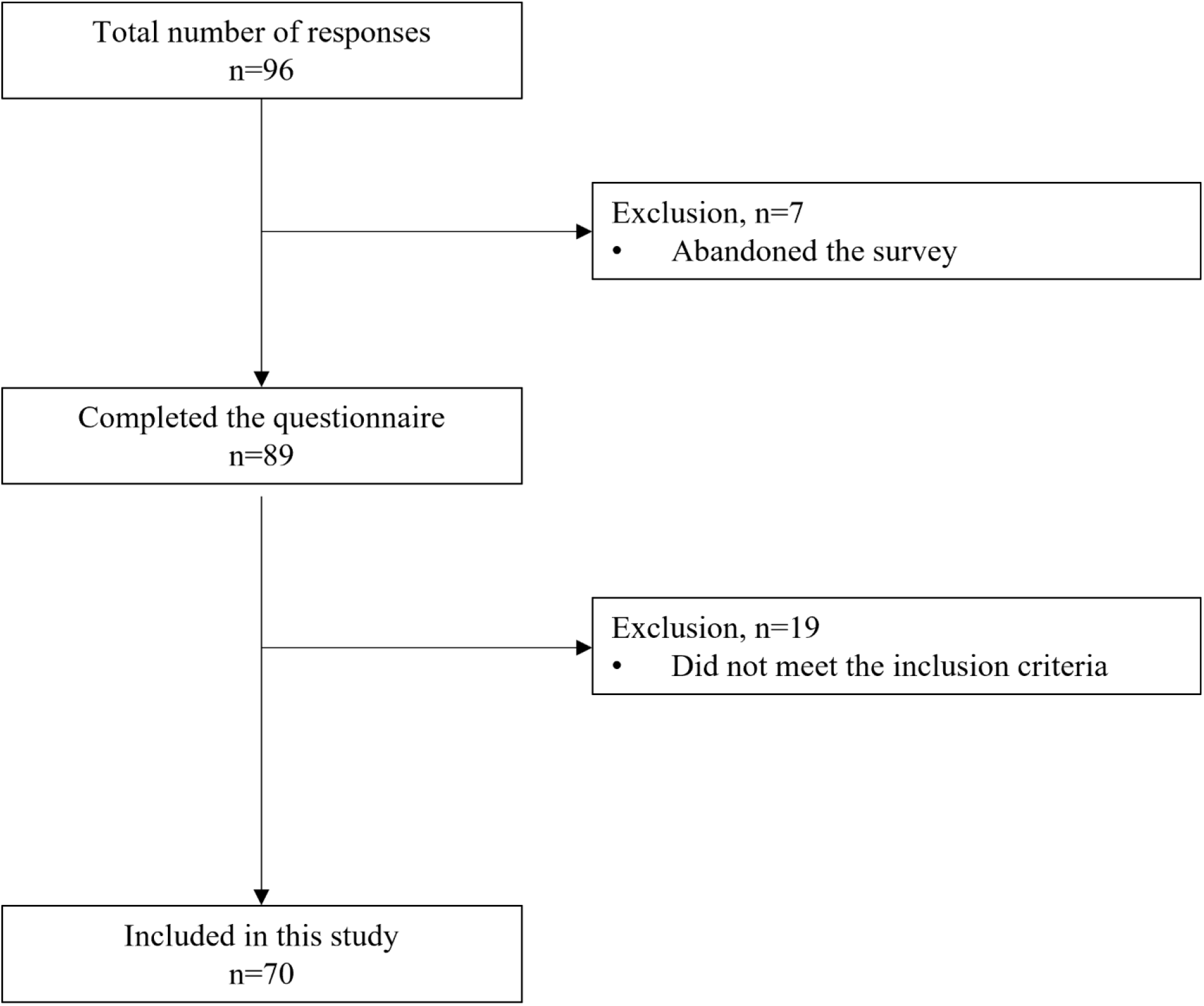
Inclusion flowchart of study participants.

### Study participants

The median age of participants was 48 years (interquartile range 42-57), and 43 (61%) were female (Table 1). Most were paediatricians (45, 64%), followed by rehabilitation medicine physicians (28, 40%), neurologists (10, 14%), and an anaesthetist (1, 1%). Only 12 (17%) participants had less than 5 years’ work experience in their specialization. The work experience in the treatment of children and adolescents with CP was similar between groups (experienced vs. unexperienced, p=0.8). Participants with experience were more likely to work in any hospital (university and/or general hospital) than unexperienced participants (42, 89% vs. 14, 61%; p=0.01). Most participants indicated that they gained their knowledge of medical cannabinoid treatment primarily through individual education (45, 64%), followed by colleagues (28, 40%), conferences (17, 24%), and as part of the education (15, 21%). 10 (14%) indicated that they had been informed by patients and families about cannabinoid therapy options.

**Table 1:** Characteristics of the participating physicians, with or without experience in treating children with cerebral palsy with cannabinoids

|  | Total<br>n=70, (%) | Experience<br>n=47, (%) | No experience<br>n=23, (%) | p-value |
| --- | --- | --- | --- | --- |
| **Sex** |  |  |  | 0.96 |
| Male | 25 (36) | 17 (36) | 8 (35) |  |
| Female | 43 (61) | 29 (62) | 14 (61) |  |
| Unknown | 2 (3) | 1 (2) | 1 (4) |  |
| **Age in years (interquartile range)** | 48 (42-57) | 47.5 (41-56) | 50 (42-60) | 0.92 |
| **Specialisation<sup>1</sup>** |  |  |  |  |
| Paediatrics | 45 (64) | 33 (70) | 12 (52) | 0.14 |
| Physical Medicine and Rehabilitation | 28 (40) | 18 (38) | 10 (44) | 0.80 |
| Neurology | 10 (14) | 8 (17) | 2 (9) | 0.35 |
| Anaesthesia | 1 (1) | 0 | 1 (4) | 0.33 |
| **Subspecialisation** |  |  |  |  |
| Yes | 53 (76) | 39 (88) | 14 (61) | 0.04 |
| Neuropediatric | 28 (40) | 19 (40) | 9 (39) | 0.92 |
| Pediatric rehabilitation medicine | 21 (30) | 18 (38) | 3 (13) | 0.05 |
| Developmental paediatrics | 8 (11) | 5 (11) | 3 (13) | 1.0 |
| Pediatric palliative care | 5 (7) | 3 (6) | 2 (9) | 1.0 |
| Pediatric oncology-haematology | 1 (1) | 1 (2) | 0 (0) | 1.0 |
| Neurometabolic | 1 (1) | 0 (0) | 1 (4) | 1.0 |
| No | 17 (24) | 8 (17) | 9 (39) |  |
| **Work experience in specialty in years** |  |  |  | 0.81 |
| 0-5 | 12 (17) | 8 (17) | 4 (32) |  |
| 6-10 | 13 (19) | 8 (17) | 5 (22) |  |
| 11-15 | 10 (14) | 8 (17) | 2 (9) |  |
| >15 | 35 (50) | 23 (49) | 12 (52) |  |
| **Work experience in the treatment of children with CP in years** |  |  |  | 0.72 |
| 0-5 | 15 (21) | 9 (19) | 6 (26) |  |
| 6-10 | 16 (23) | 11 (23) | 5 (22) |  |
| 11-15 | 18 (26) | 14 (30) | 4 (17) |  |
| >15 | 21 (30) | 13 (28) | 8 (35) |  |
| **Workplace<sup>1</sup>** |  |  |  | 0.01 |
| Any hospital | 56 (80) | 42 (89) | 14 (61) |  |
| University hospital | 23 (33) | 20 (43) | 3 (13) |  |
| General hospital | 27 (39) | 18 (38) | 9 (39) |  |
| University and general hospital | 6 (9) | 4 (8) | 2 (9) |  |
| Rehabilitation centre | 12 (17) | 4 (8) | 8 (35) |  |
| Private/Joined practice | 2 (3) | 2 (4) | 0 (0) |  |
| **Knowledge Acquisition<sup>1</sup>** |  |  |  |  |
| Individual learning | 45 (64) | 33 (70) | 12 (52) | 0.14 |
| Colleagues | 28 (40) | 19 (40) | 9 (39) | 0.92 |
| Conferences | 17 (24) | 11 (23) | 6 (26) | 0.81 |
| Part of education | 15 (21) | 10 (21) | 5 (22) | 0.97 |
| Parents & families | 10 (14) | 7 (15) | 3 (13) | 0.84 |
| Institution | 8 (11) | 6 (13) | 2 (9) | 0.62 |
| Companies | 2 (3) | 1 (2) | 1 (4) | 0.60 |
| **Country of Residence (grouped)** |  |  |  | 0.004 |
| Switzerland | 23 (33) | 15 (32) | 8 (35) |  |
| Europe excluding Switzerland | 27 (39) | 13 (28) | 14 (61) |  |
| North America | 18 (26) | 18 (38) | 0 (0) |  |
| Australia | 2 (3) | 1 (2) | 1 (4) |  |

CP, cerebral palsy
<sup>1</sup>Multiple responses possible
<sup>2</sup>Chi-square, Fisher's exact, or Wilcoxon rank-sum tests

Most participants came from Europe, followed by Switzerland, North America, and Australia (Figure 2). Characteristics of the participating physicians by region are provided in Appendix A.1.

**Figure 2:**
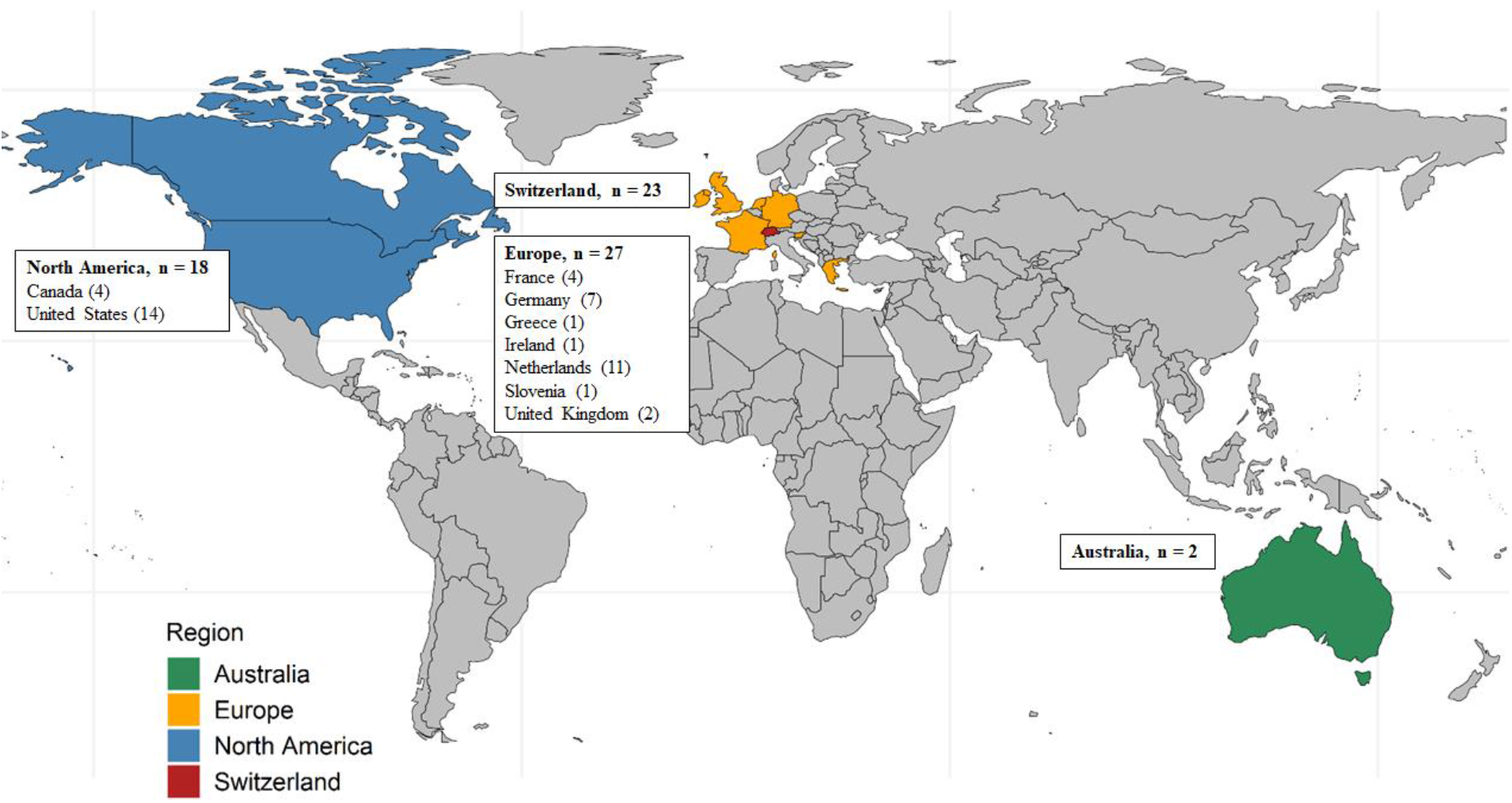
Map showing the country of residence of the participating physicians by region.

### Indications

The most frequent indication for the treatment with cannabinoid medication in children with CP was epilepsy (48, 69%), followed by spasticity (45, 64%), pain (44, 63%), behavioural problems (12, 17%), sleeping disturbance (11, 16%), and dystonia (8, 11%; Table 2). Experienced participants were more likely to indicate medical cannabinoids for epilepsy, spasticity, and a tendency towards pain compared to the unexperienced ones. Medical cannabinoids were most frequently prescribed as a co-medication (28, 40.0%) and as a second line treatment (16, 23%). The most common reasons not to initiate cannabinoids in children with CP were lack of cost coverage (24, 34%), age of the patient (19, 27%), lack of evidence on effectiveness and side effects (15, 21%), and parents wish (13,19%).

**Table 2:** Indications for treatment of children with CP with medical cannabinoids and reasons to not initiate such treatment.

|  | **Total**<br>**n=70, (%)** | **Experience**<br>**n=47, (%)** | **No experience**<br>**n=23, (%)** | **p-value<sup>3</sup>** |
| --- | --- | --- | --- | --- |
| **Indication (overall)<sup>1,2</sup>** |  |  |  |  |
| Epilepsy | 48 (69) | 37 (79) | 11 (48) | 0.009 |
| Spasticity | 45 (64) | 34 (72) | 11 (48) | 0.04 |
| Pain | 44 (63) | 33 (70) | 11 (48) | 0.06 |
| Behavioural problems | 12 (17) | 9 (19) | 3 (13) | 0.74 |
| Sleep disturbance | 11 (16) | 8 (17) | 3 (14) | 0.67 |
| Dystonia | 8 (11) | 7 (15) | 1 (4) | 0.26 |
| None | 5 (7) | 1 (2) | 4 (17) | 0.04 |
| **Context of prescription** |  |  |  | 0.01 |
| Co-medication | 28 (40) | 22 (47) | 6 (26) |  |
| Second line treatment | 16 (23) | 11 (23) | 5 (22) |  |
| Palliative treatment | 7 (10) | 6 (13) | 1 (4) |  |
| First line treatment | 2 (3) | 2 (4) | 0 (0) |  |
| Parents initiated treatment | 2 (3) | 2 (4) | 0 (0) |  |
| Not applicable | 6 (9) | 2 (4) | 4 (17) |  |
| No response | 9 (13) | 2 (4) | 7 (30) |  |
| **Reasons to not initiate cannabinoids<sup>1</sup>** |  |  |  |  |
| Lack of cost coverage | 24 (34) | 19 (40) | 5 (22) | 0.12 |
| Age of the patient | 19 (27) | 15 (32) | 4 (17) | 0.26 |
| Lack of evidence on effectiveness and side effects | 15 (21) | 10 (21) | 5 (22) | 0.97 |
| Parents wish | 13 (19) | 10 (21) | 3 (13) | 0.52 |
| Drug interaction | 10 (14) | 8 (17) | 2 (9) | 0.48 |
| Other co-morbidities | 3 (4) | 3 (6) | 0 (0) | 0.55 |
| None | 8 (11) | 5 (11) | 3 (13) | 1.00 |

<sup>1</sup>Multiple responses possible
<sup>2</sup>Most important and further indications grouped
<sup>3</sup>Chi-square, Fisher's exact, or Wilcoxon rank-sum tests

### Prescribed formulas

Participants with experience predominantly prescribed dronabinol solution (Δ9-THC 2.5%; 30%) and cannabis oil (Δ9-THC/CBD = 11/24mg per g; 17%) among the standardized formulas (Table 3). However, there was a large proportion of participants (12, 26%) who used self-medications (variable contents of Δ9-THC and CBD). Six (13%) participants did not provide specific information in response to this question. Participants without experience predominantly prescribed cannabis oil ((Δ9-THC/CBD = 11/24mg per g; 13%), CBD (9%), dronabinol solution (Δ9-THC 2.5%; 9%) and cannabis sativa spray (Δ9-THC/CBD = 2.7/2.5 mg per spray; 9%), but almost half of them (48%) did not provide information.

**Table 3:** Cannabinoids used for the treatment of children with cerebral palsy

| **Total** | **Total**<br>**n=70, (%)** | **Experience**<br>**n=47, (%)** | **No experience**<br>**n=23, (%)** |
| --- | --- | --- | --- |
| **Preparation** |  |  |  |
| Dronabinol solution: $\Delta^9$-THC 2.5% | 16 (23) | 14 (30) | 2 (9) |
| Self-medication: Diff. contents of $\Delta^9$-THC & CBD | 13 (19) | 12 (26) | 1 (4) |
| Cannabis oil: $\Delta^9$-THC/CBD = 11/24mg per g | 11 (16) | 8 (17) | 3 (13) |
| Cannabis tincture: $\Delta^9$-THC/CBD = 11/22mg per g | 3 (4) | 2 (4) | 1 (4) |
| Cannabis sativa spray (Sativex): $\Delta^9$-THC/CBD = 2.7/2.5 mg<br>per spray | 4 (6) | 2 (4) | 2 (9) |
| CBD 2,5,10% | 3 (4) | 1 (2) | 2 (9) |
| Epidiolex® | 3 (4) | 2 (4) | 1 (4) |
| No information | 17 (24) | 6 (13) | 11 (48) |

CBD, cannabidiol; THC, tetrahydrocannabinol.

### Perceived effect of the treatment and side effects

The perceived effect of the treatment with medical cannabinoids was reported to be strong or moderate by 32 (68%), weak in 10 (21%), and insignificant by 1 (2%) of the experienced participants (Table 4). Twenty-nine (41%) participants reported short-term side effects and more experienced participants than unexperienced ones reported side-effects (25, 53% vs. 4, 17%, p=0.004). The most frequently reported side effects among the experienced participants were drowsiness (12, 26%), somnolence (9, 19%), fatigue (6, 13%), and diarrhoea (6, 13%). Less frequently reported side effects are listed in Table 4. No long-term side effects were described.

**Table 4:** Short- and long-term side-effects observed in cerebral palsy children treated with cannabinoids

| **Total** | **Total**<br>**n=70, (%)** | **Experience**<br>**n=47, (%)** | **No experience**<br>**n=23, (%)** |
| --- | --- | --- | --- |
| **Effectiveness (physician reported)** |  |  |  |
| Insignificant | 2 (3) | 1 (2) | 1 (4) |
| Weak | 12 (17) | 10 (21) | 2 (9) |
| Moderate | 35 (50) | 28 (60) | 7 (30) |
| Strong | 6 (9) | 4 (9) | 2 (9) |
| No response | 15 (21) | 4 (9) | 11 (48) |
| **Short-term side effects** |  |  |  |
| Yes | 29 (41) | 25 (53) | 4 (17) |
| Drowsiness | 14 (20) | 12 (26) | 2 (9) |
| Somnolence | 10 (14) | 9 (19) | 1 (4) |
| Fatigue | 6 (9) | 6 (13) | 0 (0) |
| Diarrhoea | 6 (9) | 6 (13) | 0 (0) |
| Nausea | 5 (7) | 4 (9) | 1 (4) |
| Anxiety | 4 (6) | 4 (9) | 0 (0) |
| Confusion | 4 (6) | 4 (9) | 0 (0) |
| Disorientation | 3 (4) | 3 (6) | 0 (0) |
| Euphoria | 3 (4) | 3 (6) | 0 (0) |
| Dry mouth | 2 (3) | 2 (4) | 0 (0) |
| Hallucination | 2 (3) | 2 (4) | 0 (0) |
| Vomiting | 2 (3) | 2 (4) | 0 (0) |
| Weakness | 1 (1) | 1 (2) | 0 (0) |
| Asthenia | 1 (1) | 1 (2) | 0 (0) |
| Depression | 1 (1) | 1 (2) | 0 (0) |
| Dizziness | 1 (1) | 1 (2) | 0 (0) |
| Paranoia | 1 (1) | 1 (2) | 0 (0) |
| Psychosis | 1 (1) | 1 (2) | 0 (0) |
| Hypotonia | 1 (1) | 1 (2) | 0 (0) |
| Increased liver enzymes | 1 (1) | 1 (2) | 0 (0) |
| No | 41 (59) | 22 (47) | 19 (83) |
| **Long-term side effects** |  |  |  |
| Yes | 0 (0) | 0 (0) | 0 (0) |
| No | 41 (59) | 35 (75) | 6 (26) |

## Discussion

This international online survey assessed the prescription practices of medical cannabinoids in children with CP by paediatricians. The participating physicians (n=70) mainly acquired their knowledge on medical cannabinoids outside of medical training. The physicians frequently prescribed differing formulas of cannabinoids for various indications in children with CP. The most common indications were epilepsy, spasticity and pain, and treatment was initiated as co-medication or second line. Overall, a moderate efficacy of cannabinoids and no long-term side effects were reported.

Participants from a range of countries took part in the survey. Most participants who prescribed cannabinoids for children with CP worked in clinics, especially in university hospitals, while participants without experience in prescribing these drugs often worked in rehabilitation centres. Participants indicate that they acquired their knowledge on the topic primarily through individual learning. This highlights the need to promote knowledge transfer and education on the topic.

Medicinal cannabinoids are used for various indications in childhood and adulthood [12-14, 17-20, 22, 23, 26, 27]. In children, the evidence for most indications is limited. Nevertheless, there are numerous reports that document the use of cannabinoids for other indications [13]

In our study, epilepsy was among the three most frequently reported indications for cannabinoids in children with CP. These prescriptions are probably based on the benefit of the treatment with cannabinoids of specific forms of epilepsy in children [14-17]. However, the verification is based on the use of CBD compounds in children with Lennox-Gastaut syndrome or Dravet syndrome. Physicians seem to use cannabinoids to treat epilepsy in young CP patients with associated epilepsy based on these findings. We can not assess which forms of epilepsy were treated using medical cannabinoids and whether they were used as the sole substance or as an add-on therapy. Also, it remains unclear whether cannabinoids were used for the treatment of epilepsy or oher symptoms.

Spasticity was also among the most often reported indications for cannabinoids in our survey. Cannabinoids are used to treat spasticity in adult CP patients [18-20, 28], and the treatment of young patients is discussed repeatedly [27]. The high prescription rate of cannabinoids for spasticity in children with CP in our study could form on the evidence in adults. However, a recent RCT could not show a benefit cannabinoids in the treatment of spasticity in children with CP [24]. We run our survey before the publication of this RCT. This recent finding might reduce the prescription rate of cannabinoids to treat spasticity in children with CP in the future.

The third of the most common indications of cannabinoids in children with CP was pain. This finding was to be expected, as cannabinoids are used to treat chronic pain of various aetiologies [29, 30], and chronic pain is a common problem in children with CP, especially in more severe forms [31]. However, the efficacy of cannabinoids to relievie neuropathic pain in children is not supported [13]. Studies on the use of cannabinoids to specifically treat pain in children with CP do not exist yet. An interesting aspect of the survey was the responses regarding dystonia. Several colleagues independently indicated dystonia as their most important indication in the personal comments. To date, there is little information regarding the use of cannabinoids in dystonia. Individual case reports from the adult literature describe positive effects [32]. A recently published pilot study in children with complex movement disorders described improvements in dystonia with the use of cannabinoids [33].

Our study revealed that a variety of cannabis preparations with widely varying THC and CBD content are used in children with CP. The frequent use of cannabidiol compounds is probably due to many colleagues primarily treating patients with epilepsy. Unfortunately, we cannot determine from our survey which substance was used for which indication. In the multiple-choice question addressing the prescribed preparations, we only listed the preparations that are best known to us and most frequently used in Switzerland. However, many colleagues stated that they used other preparations. The reasons for the choice of preparations were not asked. We also do not know which preparations are predominantly available in which countries and on what legal basis they can be acquired. The pharmacologically manufactured Nabiximols (Sativex®), which can be purchased as standard in many countries, was used rarely.

The perceived effect of medical cannabinoids was reported to be moderate by most experienced physicians. However, the survey does not allow conclusions to be drawn about the effectiveness of cannabinoids for various indications. Half of the experienced physicians described short-term side effects, drowsiness and somnolence were most common. No long-term adverse effects were perceived. Nonetheless the survey was not designed to cover the safety spectrum of medicinal cannabinoids. Such a survey is not capable of doing this, nor do we wish to make a definitive statement about the safety. Still, our findings on side effects is in line with previous research [34, 35]. To answer questions on efficacy and safety of medical cannabinoids in children with CP, interventional studies with a randomized controlled design are mandatory.

This study has some limitations. The survey format provides insight into the topic and an impression of whether and how colleagues in western countries treat children with CP with cannabinoids. However, our observation is not representative. We cannot determine the response rate as we used several distribution paths. Statements about national preferences are not possible as we obtained a convenience sample mainly driven by personal contacts.

As it is not possible to track how many physicians received the survey and how many completed it, we could not assess the response rate. It is possible, for example, that colleagues who felt concerned by the topic completed the survey, biasing our findings. Also, the number of participants per region is too small to draw conclusions per region. Furthermore, it is not possible to check the quality of the data, as the survey was conducted anonymously. Accordingly, the results of our study should be interpreted with caution and no conclusions must be drawn regarding the safety and efficacy of cannabinoids in children with CP.

## Conclusions

This survey shows that medical cannabinoids are prescribed for a wide range of indications in children with CP in western countries. Despite the lack of evidence for their use in this patient group. Accordingly, it is important that further RCTs clarify for which indications and in which situations the use of cannabinoids is justified. Regarding epilepsy, it seems important to closely examine the use of cannabidiol specifically in CP patients. Further studies should also include indications, such as pain, behavioural problems, or dystonia. This will require multicentre RCTs, which could be based, for example, within national or international research platforms such as the Swiss Cerebral Palsy Registry.

## Data Availability

All data produced in the present study are available upon reasonable request to the authors

CBD: cannabidiol
CP: Cerebral palsy
RCT: randomized controlled trial
THC: tetrahydrocannabidiol;

## Acknowledgements

We would like to thank all physicians who took the time to complete our survey and who shared it with colleagues. Without them, this work would not have been possible. We would also like to thank all members of the Swiss Cerebral Palsy Registry who gave the input for this survey with their ideas and comments on the topic.

## Appendices

### Appendix A: Additional table

**Table A.1:** Characteristic of the participating physicians by region.

|  | Total<br>n=70, (%) | Switzerland<br>n=23, (%) | Europe<br>excluding<br>Switzerland<br>n=27, (%) | North<br>America<br>n=18, (%) | Australia<br>n=2, (%) | p-value<sup>3,4</sup> |
| --- | --- | --- | --- | --- | --- | --- |
| **Sex** |  |  |  |  |  | 0.07 |
| Male | 25 (36) | 0 (0) | 5 (19) | 0 (0) | 0 (0) |  |
| Female | 43 (61) | 11 (48) | 20 (74) | 10 (56) | 2 (100) |  |
| Unknown | 2 (3) | 12 (52) | 2 (4) | 8 (44) | 0 (0) |  |
| **Age in years (interquartile range)** | 48 (42-57) | 49 (46-57) | 51 (42-61) | 41.5 (38-55) | 40 (38-42) |  |
| **Workplace<sup>1</sup>** |  |  |  |  |  | <0.001 |
| Any hospital | 56 (80) | 20 (87) | 16 (59) | 17 (94) | 2 (100) |  |
| University hospital | 23 (33) | 11 (48) | 5 (19) | 7 (39) | 0 (0) |  |
| General hospital | 27 (39) | 9 (39) | 10 (37) | 6 (33) | 2 (100) |  |
| University and general hospital | 6 (9) | 1 (4) | 1 (4) | 4 (22) | 0 (0) |  |
| Rehabilitation centre | 12 (17) | 0 (0) | 11 (41) | 1 (6) | 0 (0) |  |
| Private/Joined practice | 2 (3) | 2 (9) | 0 (0) | 0 (0) | 0 (0) |  |
| **Work experience with cannabinoids in years** |  |  |  |  |  | 0.80 |
| 0-5 | 64 (91) | 21 (91) | 25 (93) | 16 (89) | 0 (0) |  |
| 6-10 | 5 (7) | 2 (9) | 2 (7) | 1 (6) | 2 (100) |  |
| 11-15 | 1 (1) | 0 (0) | 0 (0) | 1 (6) | 0 (0) |  |
| **Indication (overall)<sup>1,2</sup>** |  |  |  |  |  |  |
| Epilepsy | 48 (69) | 17 (74) | 13 (48) | 16 (89) | 2 (100) | 0.01 |
| Spasticity | 45 (64) | 19 (83) | 15 (56) | 10 (56) | 1 (50) | 0.08 |
| Pain | 44 (63) | 19 (83) | 14 (52) | 10 (56) | 1 (50) | 0.06 |
| Behavioural problems | 12 (17) | 5 (22) | 5 (19) | 1 (6) | 1 (50) | 0.41 |
| Sleep disturbance | 11 (16) | 5 (22) | 3 (11) | 2 (11) | 1 (50) | 0.57 |
| Dystonia | 8 (11) | 2 (9) | 4 (15) | 2 (11) | 0 (0) | 0.90 |
| None | 5 (7) | 1 (4) | 4 (15) | 0 (0) | 0 (0) | 0.21 |
| **Context of prescription** |  |  |  |  |  | 0.06 |
| Co-medication | 28 (40) | 10 (43.5) | 7 (25.9) | 10 (55.6) | 1 (50.0) |  |
| Second line treatment | 16 (23) | 6 (26.1) | 4 (14.8) | 4 (22.2) | 0 (0.0) |  |
| Palliative treatment | 7 (10) | 5 (21.7) | 2 (7.4) | 0 (0.0) | 0 (0.0) |  |
| First line treatment | 2 (3) | 0 (0.0) | 1 (3.7) | 1 (5.6) | 0 (0.0) |  |
| Parents initiated treatment | 2 (3) | 0 (0.0) | 2 (7.4) | 0 (0.0) | 0 (0.0) |  |
| Not applicable | 6 (9) | 1 (4.4) | 4 (14.8) | 1 (5.6) | 0 (0.0) |  |
| No response | 9 (13) | 1 (4.4) | 7 (25.9) | 1 (5.6) | 0 (0.0) |  |
| **Reasons for not initiating cannabinoids<sup>1</sup>** |  |  |  |  |  |  |
| Lack of cost coverage | 24 (34) | 5 (22) | 10 (37) | 9 (50.0) | 0 (0.0) | 0.18 |
| Age of the patient | 19 (27) | 8 (35) | 4 (15) | 7 (38.9) | 0 (0.0) | 0.12 |
| Lack of evidence on effectiveness and side effects | 15 (21) | 4 (17) | 8 (30) | 3 (16.7) | 0 (0.0) | 0.57 |
| Parents wish | 13 (19) | 4 (17) | 4 (15) | 5 (28) | 0 (0) | 0.59 |
| Drug interaction | 10 (14) | 1 (4) | 4 (15) | 5 (28) | 0 (0) | 0.13 |
| Other co-morbidities | 3 (4) | 0 (0) | 1 (4) | 2 (11) | 0 (0) | 0.26 |
| None | 8 (11) | 6 (26) | 1 (4) | 1 (6) | 0 (0) | 0.43 |

<sup>1</sup>Multiple responses possible
<sup>2</sup>Most important and further indications grouped
<sup>3</sup>Excluding Australia due to low sample size.
<sup>4</sup>Chi-square, Fisher's exact, or Wilcoxon rank-sum tests

### Appendix B: Online survey

#### Appendix B.1: Questionnaire

Q1 Have you already worked with patients treated with cannabinoid drugs?
  □ *Yes*
  □ *No*
Q2 Were any of these in a paediatric age (0-18 years)?
  □ *Yes*
  □ *No*
Q3 Were any of these children/adolescents diagnosed with cerebral palsy?
  □ *Yes*
  □ *No*
Q4 Did you ever experience a situation in which the prescription of medical cannabinoids in this type of patients was debated, but for some reason not prescribed?
  □ *Yes*
  □ *No*
Q5 Before we continue with the survey, please inform us how you answered the first 4 questions.
  □ *(If at least one of the questions Q1-Q4 was answered affirmatively, access to Q6-Q34 was provided*.)
  □ *I answered the first four questions all with no*
  □ *I answered at least one of the first four questions with a yes*
Q6 What is your country of birth? _____________________________
Q7 In what year were you born? (Enter 4-digit birth year; for example, 1976) _____________________________
Q8 What is your gender?
  □ *Male*
  □ *Female*
Q9 Where did you attain your medical degree (enter city and country)? _____________________________
Q10 What is your medical specialization?
  □ *Paediatrics*
  □ *Neurology*
  □ *Neurosurgery*
  □ *Child and adolescent psychiatry and psychotherapy*
  □ *Anaesthesia*
  □ *Medical genetics*
  □ *Psychiatry and psychotherapy*
  □ *Radiology*
  □ *Physical medicine and rehabilitation*
  □ *General internal medicine*
  □ *Orthopaedics*
  □ *Paediatric surgery*
  □ *Other (Specify)*
Q11 How many years of experience do you have in your specialty? (If more than one specialty, please specify the numbers for every single one, in the comment box below.)
  □ *0-5*
  □ *6-10*
  □ *11-15*
  □ *>15*
Q12 Have you also attained a paediatric subspecialty?
  □ *Yes*
  □ *No*
Q13 Please specify your subspecialty.
  □ *Developmental paediatrics*
  □ *Paediatric rehabilitation medicine*
  □ *Paediatric palliative care*
  □ *Neuropediatrics*
  □ *Paediatric oncology-haematology*
  □ *Other (specify)*
Q14 How many years of experience do you have in your subspecialty? (If more than one subspecialty, please specify the numbers for every single one, in the comment box below.)
  □ *0-5*
  □ *6-10*
  □ *11-15*
  □ *>15*
Q15 In which country do you currently work? _____________________________
Q16 What kind of institution do you currently work in?
  □ *Hospital*
  □ *Private practice*
  □ *Joined practice*
  □ *University hospital*
  □ *Other (specify)*
Q17 How were you introduced to medical cannabinoids?
  □ *As part of my education*
  □ *Participation in a congress on the theme*
  □ *Individual learning on the subject*
  □ *Through the advice of other collaborators*
  □ *Through education provided by my institution*
  □ *Through education provided by pharmaceutical companies*
  □ *Other (specify)*
Q18 How many years have you been treating children diagnosed with cerebral palsy?
  □ *0-5*
  □ *6-10*
  □ *11-15*
  □ *16-20*
  □ *>20*
Q19 How many of your patients diagnosed with cerebral palsy are/were treated with medical cannabinoids? ___________________________________________________________________________
Q20 How long have you been working with cannabinoids for therapeutic purposes? (years)
  □ *0-5*
  □ *6-10*
  □ *11-15*
  □ *16-20*
  □ *>20*
Q21 What is, in your experience, the most important indication to start a treatment with cannabinoids in children with cerebral palsy?
  □ *Pain*
  □ *Epilepsy*
  □ *Sleep disturbance*
  □ *Spasticity*
  □ *Behavioural problems*
  □ *None*
  □ *Other (specify)*
Q22 How would you assess, on a scale from 1 to 5, the effectiveness of the therapy in the context of the indication above? (Scale values are specified below.)
  □ *Insignificant*
  □ *Weak*
  □ *Moderate*
  □ *Strong*
  □ *Excellent*
Q23 Would you also consider further indications?
  □ *Pain*
  □ *Spasticity*
  □ *Epilepsy*
  □ *Behavioural problems*
  □ *Sleep disturbance*
  □ *No*
  □ *Other (specify)*
Q24 In which context was the therapy mainly started?
  □ *First line treatment*
  □ *Second line treatment*
  □ *Co-medication*
  □ *Palliative treatment*
  □ *Other (specify)*
Q25 Have there been any criteria that prevented/suggested not to undertake this therapy in paediatric patients?
  □ *Age of the patient*
  □ *Parents’ wishes*
  □ *Other pathologies (contraindications)*
  □ *Lack of cost coverage by the health insurance*
  □ *Legal or administrative hurdles*
  □ *Other medicines (drug interactions)*
  □ *None*
  □ *Other (specify)*
Q26 Which type of drug has mostly been used? (The list below only lists the preparations offered in Switzerland, if you have prescribed other preparations, please specify below.)
  □ *Cannabis sativa spray (Sativex): viscous extract of Δ9-THC - and CBD, standardized contents: 2*.*7/2*.*5 mg per spray*
  □ *Dronabinol solution: synthetically produced Δ9-THC, standardized dronabinol content 2,5%*
  □ *Cannabis tincture: alcoholic drop solution, standardized Δ9-THC and CBD contents: 11/22mg per g*
  □ *Cannabis oil: oily drop solution, standardized Δ9-THC and CBD contents: 11/24mg per g*
  □ *Sativa oil: oily drop solution, standardized Δ9-THC and CBD contents (Sativex equivalent ratio*
  □ *2,7/2,5 mg per g)*
  □ *Self-medication: different contents of Δ9-THC, CBD, and other cannabinoids*
  □ *Other (specify)*
Q27 What was the maximum dosage prescribed for the respective preparations? (Please enter how it was prescribed and the dose of THC and/or CDB in mg/kg/day.) ___________________________________________________________________________
Q28 Was there a habituation effect noticed?
  □ *Yes*
  □ *No*
Q29 Was a dose adjustment necessary as a result?
  □ *Yes*
  □ *No*
Q30 Were any important short-term adverse events noticed?
  □ *Anxiety*
  □ *Asthenia*
  □ *Balance*
  □ *Confusion*
  □ *Depression*
  □ *Diarrhoea*
  □ *Disorientation*
  □ *Dizziness*
  □ *Dry mouth*
  □ *Dyspnoea*
  □ *Euphoria*
  □ *Eye disorders*
  □ *Fatigue*
  □ *Hallucination*
  □ *Nausea*
  □ *Paranoia*
  □ *Psychosis*
  □ *Seizures*
  □ *Somnolence*
  □ *Vomiting*
  □ *Weakness*
  □ *None*
  □ *Other (specify)*
Q31 What consequences have they had for the continuation of the therapy?
  □ *Irrelevant*
  □ *Drug dose adjustment*
  □ *Therapy stop*
  □ *Other (specify)*
Q32 Were any important long-term adverse events noticed?
  □ *Cardiovascular disease*
  □ *Respiratory disease*
  □ *Cancer*
  □ *Psychotic disorders*
  □ *Suicide or suicidal thoughts*
  □ *None*
  □ *Other (specify)*
Q33 On a scale from 1 to 5, please give us below a personal rating for your experience with the use of these therapies in children with cerebral palsy.
  □ *Bad*
  □ *Acceptable*
  □ *Average*
  □ *Good*
  □ *Outstanding*
Q34 Remarks. ___________________

